# Causal relationship between mental disorders and abdominal aortic aneurysm: insights from the genetic perspective

**DOI:** 10.1101/2024.02.08.24302548

**Authors:** Ming-Gang Deng, Chen Chai, Kai Wang, Zhi-Hui Zhao, Jia-Qi Nie, Fang Liu, Yuehui Liang, Jiewei Liu

## Abstract

**Objective:** This study investigated the genetic link between mental disorders—depression, schizophrenia (SCZ), and bipolar disorder (BIP)—and abdominal aortic aneurysm (AAA).

**Methods:** We used global/local genetic correlation analyses and identified shared genomic loci. Bidirectional univariable Mendelian Randomization (UMR) assessed causal directions, with multivariable MR (MVMR) refining the analysis. Mediation analyses examined if known AAA risk factors influenced this relationship.

**Results:** Global correlations showed positive links between depression, SCZ, and AAA, but not BIP. Local analyses identified specific genomic regions of correlation. We found 26, 141, and 10 shared loci for AAA with depression, SCZ, and BIP, respectively. UMR indicated significant associations between genetically predicted depression (OR 1.270; 95% CI 1.071-1.504; *p* = 0.006) and SCZ (OR 1.047; 95% CI 1.010-1.084; *p* = 0.011) with AAA, but not BIP. These results were confirmed by MVMR analyses. Mediation analyses showed that smoking, hypertension, hyperlipidemia, and coronary atherosclerosis mediated the impact of depression on AAA while smoking mediated SCZ’s impact.

**Conclusion:** This study provides evidence that genetically predicted depression and SCZ are linked to an increased risk of AAA, mediated by traditional AAA risk factors.

## 1. Introduction

Abdominal aortic aneurysm (AAA) is a prevalent vascular disease marked by localized dilation of the abdominal aorta and is linked to an approximately 90% mortality risk due to rupture^1^. While recent open surgical repair and endovascular stent graft therapies have notably enhanced the treatment of AAA, there is currently no approved and effective pharmacotherapy to restrain and prevent AAA progression and rupture^2,3^. Consequently, primary preventive measures targeting the risk factors of AAA are necessary to reduce its incidence and adverse effects on health.

Mental disorders, such as depression, schizophrenia (SCZ), and bipolar disorder (BIP), have been found to lead to various adverse outcomes. For example, our previous studies revealed that individuals with depression or SCZ were more likely to be frail^4,5^. In addition, current evidence suggests that mental disorders are linked to higher risks of cardiovascular disease, which include but are not limited to heart failure, coronary artery disease, and hypertension^6–8^. Therefore, investigating the association between mental disorders and AAA is of vital importance for public health.

To our knowledge, the associations of AAA with SCZ and BIP are currently unclear, as no studies have reported them. In the case of depression and AAA, a prospective study based on the Korean National Health Insurance System database revealed that patients with AAA were more likely to be depressed^9^. Conversely, depressed individuals were found to have a higher risk of incident AAA in the Norwegian Nord-Trøndelag Health Study^10^. Despite this, the causative association between depression and AAA needs further investigation, as observational studies were susceptible to confounding bias, reverse causality, etc.

Recent large-scale genome-wide association studies (GWASs) have identified numerous genetic loci associated with depression^11^, SCZ^12^, BIP^13^, and AAA^14^. These findings enhance the statistical power and offer the opportunity to explore their genetic associations. In addition, Mendelian Randomization (MR), which uses genetic variants (single nucleotide polymorphisms, SNPs) as instruments, is a powerful method for obtaining robust causal inference^15^ and has been widely utilized in recent epidemiological research^16–18^. As a result, this study aims to investigate the genetic correlation between AAA and mental disorders and to utilize the MR design to evaluate their causality.

## 2. Methods and materials

### 2.1. Study design

An overview of this research is presented in **Fig 1**. Initially, we explored the genetic association between AAA and mental disorders from three perspectives: global genetic correlation, local genetic correlation, and identification of shared genomic loci. Following that, we utilized a bidirectional MR design to evaluate the causal association between AAA and mental disorders. Furthermore, mediation MR analyses were performed to ascertain whether the causality was mediated by known risk factors.

**Fig 1.**
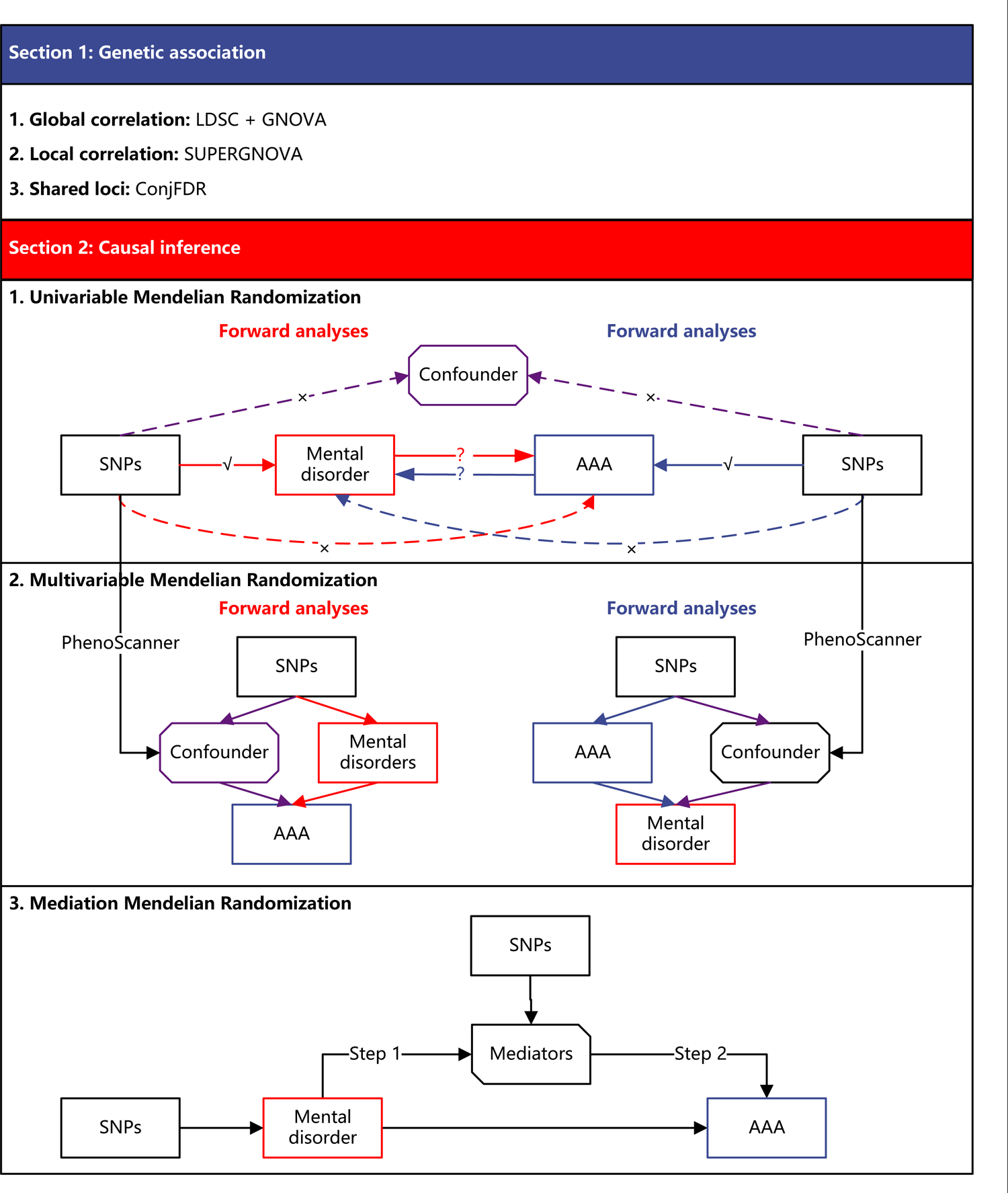
An overview of study design.

### 2.2. Data sources and ethical approval

The data sets utilized in this research were all publicly available from the previous studies, and the details were described in the **Supplementary Methods**. The ethical approvals were all obtained by the original papers, and thus the ethical approval for our study could be waived.

### 2.3. Genetic association

#### 2.3.1. Global genetic correlation

We conducted a global genetic correlation analysis to estimate the correlation between the GWAS summary statistics of AAA and three mental disorders. Two different software tools were utilized to perform this analysis. The first one is Linkage disequilibrium score regression (LDSC, https://github.com/bulik/ldsc)^19^. We used the cross-trait LDSC which only requires the GWAS summary statistics that account for sample overlap. Cross-trait LDSC analysis requires the GWAS summary statistics to be from the same ethnic group. Since the GWAS summary statistics used in this study are largely from Europeans, we used the 1000 Genomes Project European samples as the reference sample for LD score calculation.

Another complementary method for estimating global genetic correlation is the Genetic Covariance Analyzer (GNOVA)^20^. GNOVA (available at https://github.com/xtonyjiang/GNOVA) also requires only the GWAS summary statistics as input to perform global genetic correlation analysis. The major advantage of GNOVA is its ability to include omic datasets (e.g., transcriptomic and epigenomic) in the GNOVA model. The LD score was also calculated using the 1000 Genomes European samples.

#### 2.3.2. Local genetic correlation

In addition to estimating the global genetic correlation between the GWAS summary statistics of AAA and mental disorders, we also attempted to estimate the local genetic correlation between them in specific genomic regions. To perform this analysis, we used SUPERGNOVA (available at https://github.com/qlu-lab/SUPERGNOVA). SUPERGNOVA was developed within the previous GNOVA statistical framework, which includes functional annotation when estimating genetic correlation^21^. Similar to GNOVA, only GWAS summary statistics and an LD calculation reference panel were needed as input to run SUPERGNOVA.

#### 2.3.3. Shared genomic loci

To enhance the statistical power of detecting loci associated with both AAA and mental disorders, we utilized the conjunctional false discovery rate (conjFDR) method for this analysis. This method is based on standard FDR analysis and aims to identify SNPs that are associated with both target traits (e.g., SNPs that are jointly associated with AAA and SCZ). The conjFDR analysis is implemented in the pleioFDR software (available at https://github.com/precimed/pleiofdr), and it runs in the MATLAB environment^22^. The analysis followed the instructions of the pleioFDR tool with default parameters.

### 2.4. Causal inference

#### 2.4.1. Univariate Mendelian Randomization

We initially performed the univariate Mendelian Randomization (UMR) to assess the causal association between mental disorders and AAA. The statistical analyses were conducted in two directions: 1) mental disorders as the exposures to check the effect of mental disorders on AAA, and 2) AAA as exposures to evaluate whether people with AAA had a likelihood to develop mental disorders.

Independent SNPs (*R*^2^ < 0.001, window size = 10MB) were firstly screened at the genome-wide significance threshold (*p* < 5×10^-8^) to ensure their robust association with exposure and freedom from linkage disequilibrium. Subsequently, the F statistic for each SNP was calculated using the formula beta^2^/se^2^, and those with an F < 10 were considered weak instruments and were excluded. The remaining SNPs were harmonized with the outcome data, and the palindromic genetic variants were eliminated at this stage. If any genetic variants were unavailable in the outcome data, they will be replaced by the proxied SNPs if possible. After the aforementioned procedures, the remaining SNPs were used for causal inference.

The inverse variance weighted (IVW) method was used as the primary approach to obtain the causal estimate, assuming the absence of average pleiotropic effects, and in this scenario, it is the most efficient method. Cochran’s Q statistic was computed to assess the heterogeneity among the SNPs. If significant heterogeneity was detected (*p* < 0.05), the IVW method would be applied under a random-effects model; otherwise, it would be applied under a fixed-effects model. Alongside the IVW method, several other approaches, such as MR-Egger, weighted median, MR-cML (constrained maximum likelihood), and MR-RAPS (robust adjusted profile score), were employed to evaluate the robustness of the findings. In the meantime, the MR-Egger regression intercept term was utilized to evaluate the presence of horizontal pleiotropy among the instruments.

#### 2.4.2. Multivariate Mendelian Randomization

The multivariate Mendelian Randomization (MVMR), which employs SNPs associated with various and potentially related exposures, is an extension of UMR that can detect the causal effects of many risk factors simultaneously^23^. Considering that the instrumental variables in the UMR may meanwhile be linked to secondary phenotypes and even to the risk factor of outcomes, which may violate the critical assumption of MR, we further carried out the MVMR to ascertain if the UMR estimates were affected by any confounders. The PhenoScanner V2, an expanded tool for searching human genotype-phenotype associations, was employed in advance to screen the secondary phenotypes with which the instrumental variables were associated^24^. The significant confounding phenotype (*p* <1×10^-5^) would then be included as a covariate in the MVMR model. Similar to the UMR analyses, the multivariable-adjusted effect sizes were primarily estimated by the IVW method, and supplemented by the MR-Egger, weighted median, and MR-cML methods.

#### 2.4.3. Mediation effect of known risk factors

After establishing the causal relationship between mental disorders and AAA, our objective is to delve deeper into whether this causality is mediated by known risk factors. For example, smoking, hypertension, hyperlipidemia, and coronary atherosclerosis (CA) have been identified as risk factors for AAA^3^. To assess their mediation role, a two-step UMR analysis was performed. The first step involved estimating the causal effects of mental disorders on smoking, hypertension, hyperlipidemia, and coronary atherosclerosis, and the second step involved estimating the causal effects of smoking, hypertension, hyperlipidemia, and CA on AAA. The mediation proportion was calculated based on the causal effect estimated by the IVW method, while other methods, including MR-Egger, weighted median, MR-cML, and MR-RAPS, were employed as sensitivity analyses.

## 3. Results

### 3.1. Genetic association

#### 3.1.1. Global genetic correlation

The estimates of the global genetic correlation between mental disorders and AAA are presented in **Fig 2**. The LDSC method indicated that genetic variants related to depression [*r*_g_= 0.157, se = 0.023, *p* = 3.52×10^−12^] and SCZ (*r*_g_= 0.042, se = 0.020, *p* = 0.042) were positively correlated with AAA, while BIP was not (*r*_g_= 0.013, se = 0.023, *p* = 0.591). Additionally, the GNOVA method estimates further supported these findings, indicating a positive association of AAA with depression (*r*_g_= 0.152, se = 0.019, *p* = 8.41×^-16^) and SCZ (*r*_g_= 0.053, se = 0.017, *p* = 0.002). Conversely, BIP did not demonstrate a significant association with AAA (*r*_g_= 0.021, se = 0.019, *p* = 0.272).

**Fig 2.**
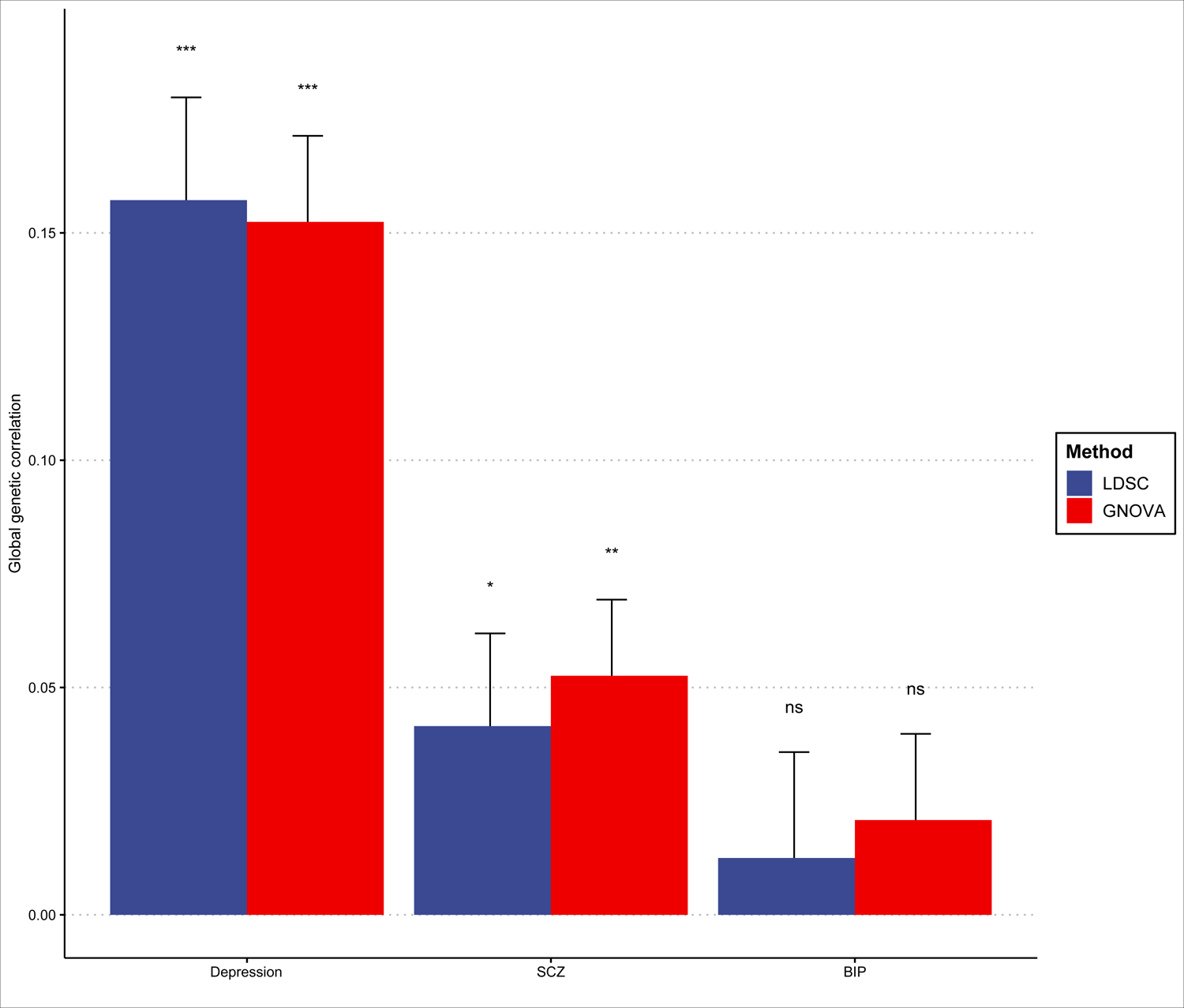
Global genetic correlation between mental disorders and abdominal aortic aneurysm. LDSC, linkage disequilibrium score regression; GNOVA, GeNetic cOVariance Analyzer; SCZ, schizophrenia; BIP, bipolar disorder; *** indicates *p* <0.001; ** indicates *p* < 0.01; * indicates *p* < 0.05; ^ns^ indicates no significance.

#### 3.1.2. Local genetic correlation

The local genetic correlation analyses revealed that mental disorders exhibited local correlations with AAA in various genomic regions, as detailed in **Table 1**. Following correction for multiple tests, depression was identified to have a positive association with AAA in four genomic regions (chr9: 126927204-128926989, chr10: 106139294-107874146, chr11: 42244050-42854352, and chr18: 77151867-78017074). Additionally, SCZ demonstrated a positive correlation with AAA in 9 out of 10 significant regions. While the global correlation did not indicate an association between BIP and AAA, the local analyses revealed correlations in 7 genomic regions, with 6 of them also showing a positive association.

**Table 1.** Locally correlated genomic regions between mental disorders and abdominal aortic aneurysm.

| Trait1&Trait2 | chr | start | end | rho | corr | h<sup>2</sup>_1 | h<sup>2</sup>_2 | var | m | p-value | p.adjust |
| --- | --- | --- | --- | --- | --- | --- | --- | --- | --- | --- | --- |
| Depression & AAA | 9 | 126927204 | 128926989 | 7.98E-05 | 0.945 | 7.26E-05 | 9.81E-05 | 2.47E-10 | 644 | 3.78E-07 | 8.57E-04 |
| Depression & AAA | 10 | 106139294 | 107874146 | 6.17E-05 | 1.112 | 1.40E-05 | 2.19E-04 | 1.72E-10 | 758 | 2.58E-06 | 2.93E-03 |
| Depression & AAA | 11 | 42244050 | 42854352 | 4.31E-05 | 1.130 | 2.05E-05 | 7.07E-05 | 9.28E-11 | 277 | 7.83E-06 | 5.92E-03 |
| Depression & AAA | 18 | 77151867 | 78017074 | 4.62E-05 | 1.159 | 2.74E-05 | 5.79E-05 | 1.12E-10 | 323 | 1.30E-05 | 7.37E-03 |
| SCZ & AAA | 2 | 73174848 | 74121000 | 7.41E-05 | 0.907 | 1.61E-05 | 4.15E-04 | 3.73E-10 | 277 | 1.24E-04 | 2.84E-02 |
| SCZ & AAA | 2 | 232791948 | 233964632 | -1.38E-04 | -1.021 | 2.56E-05 | 7.19E-04 | 6.93E-10 | 514 | 1.44E-07 | 1.64E-04 |
| SCZ & AAA | 5 | 73508509 | 75240469 | 1.21E-04 | 0.792 | 9.10E-05 | 2.57E-04 | 9.36E-10 | 766 | 7.55E-05 | 2.46E-02 |
| SCZ & AAA | 10 | 106139294 | 107874146 | 1.16E-04 | 1.355 | 1.40E-05 | 5.21E-04 | 7.46E-10 | 759 | 2.28E-05 | 1.30E-02 |
| SCZ & AAA | 10 | 130282817 | 131192968 | 1.08E-04 | 1.746 | 3.02E-05 | 1.27E-04 | 7.75E-10 | 576 | 1.02E-04 | 2.59E-02 |
| SCZ & AAA | 11 | 16790799 | 18788126 | 1.76E-04 | 0.722 | 6.85E-05 | 8.64E-04 | 1.90E-09 | 974 | 5.51E-05 | 2.19E-02 |
| SCZ & AAA | 12 | 2176265 | 2886299 | 9.31E-05 | 0.815 | 6.39E-06 | 2.04E-03 | 5.36E-10 | 367 | 5.76E-05 | 2.19E-02 |
| SCZ & AAA | 14 | 102341650 | 104759919 | 2.23E-04 | 0.671 | 8.11E-05 | 1.37E-03 | 2.12E-09 | 801 | 1.28E-06 | 9.75E-04 |
| SCZ & AAA | 15 | 78640760 | 79259009 | 2.80E-04 | 0.777 | 1.88E-04 | 6.91E-04 | 1.27E-09 | 202 | 3.75E-15 | 8.56E-12 |
| SCZ & AAA | 17 | 820511 | 1480393 | 1.05E-04 | 1.043 | 2.83E-05 | 3.56E-04 | 7.27E-10 | 264 | 1.01E-04 | 2.59E-02 |
| BIP & AAA | 2 | 73174848 | 74121000 | 8.47E-05 | 1.036 | 1.61E-05 | 4.15E-04 | 4.64E-10 | 277 | 8.37E-05 | 3.01E-02 |
| BIP & AAA | 8 | 24457122 | 25461315 | -2.03E-04 | -0.944 | 7.38E-05 | 6.29E-04 | 1.71E-09 | 466 | 9.08E-07 | 1.04E-03 |
| BIP & AAA | 12 | 47505424 | 48267360 | 1.90E-04 | 1.252 | 5.57E-05 | 4.15E-04 | 1.27E-09 | 379 | 9.37E-08 | 2.14E-04 |
| BIP & AAA | 15 | 66948951 | 68640917 | 2.27E-04 | 1.046 | 9.96E-05 | 4.72E-04 | 2.86E-09 | 789 | 2.18E-05 | 1.10E-02 |
| BIP & AAA | 15 | 78640760 | 79259009 | 1.71E-04 | 0.873 | 1.88E-04 | 2.03E-04 | 1.63E-09 | 202 | 2.41E-05 | 1.10E-02 |
| BIP & AAA | 19 | 10030690 | 11279257 | 2.63E-04 | 0.585 | 1.35E-04 | 1.49E-03 | 4.52E-09 | 482 | 9.23E-05 | 3.01E-02 |
| BIP & AAA | 22 | 43187900 | 43645335 | 1.21E-04 | 1.231 | 1.47E-05 | 6.53E-04 | 8.14E-10 | 257 | 2.31E-05 | 1.10E-02 |

**Abbreviations:** SCZ, schizophrenia; AAA, abdominal aortic aneurysm; BIP, bipolar disorder; chr, chromosome; start/end, the start or end position of the genomic region from the input genome partition file; rho, the estimation of local genetic covariance; corr, the estimation of local genetic correlation; h<sup>2</sup>\_1/h<sup>2</sup>\_2, The estimation of local heritability of trait 1/2; var, the variance of the estimation of local genetic covariance; m, the number of SNPs involved in the estimation of local genetic covariance in the genomic region.

#### 3.1.3. Shred genetic variants

In line with the preceding analyses, a multitude of shared genetic variants have been discerned between mental disorders and AAA. The findings are meticulously outlined in **Fig. 3** and **Table S1**. Specifically, there are 26 independent SNPs shared between depression and AAA, with 21 of them demonstrating a consistent directional effect on both conditions. Similarly, SCZ and AAA exhibit the sharing of 141 independent loci, among which 21 loci manifest an opposing effect. Notably, only 10 genetic variants are concurrently associated with BIP and AAA, with 4 of them displaying the same directional effect.

**Fig 3.**
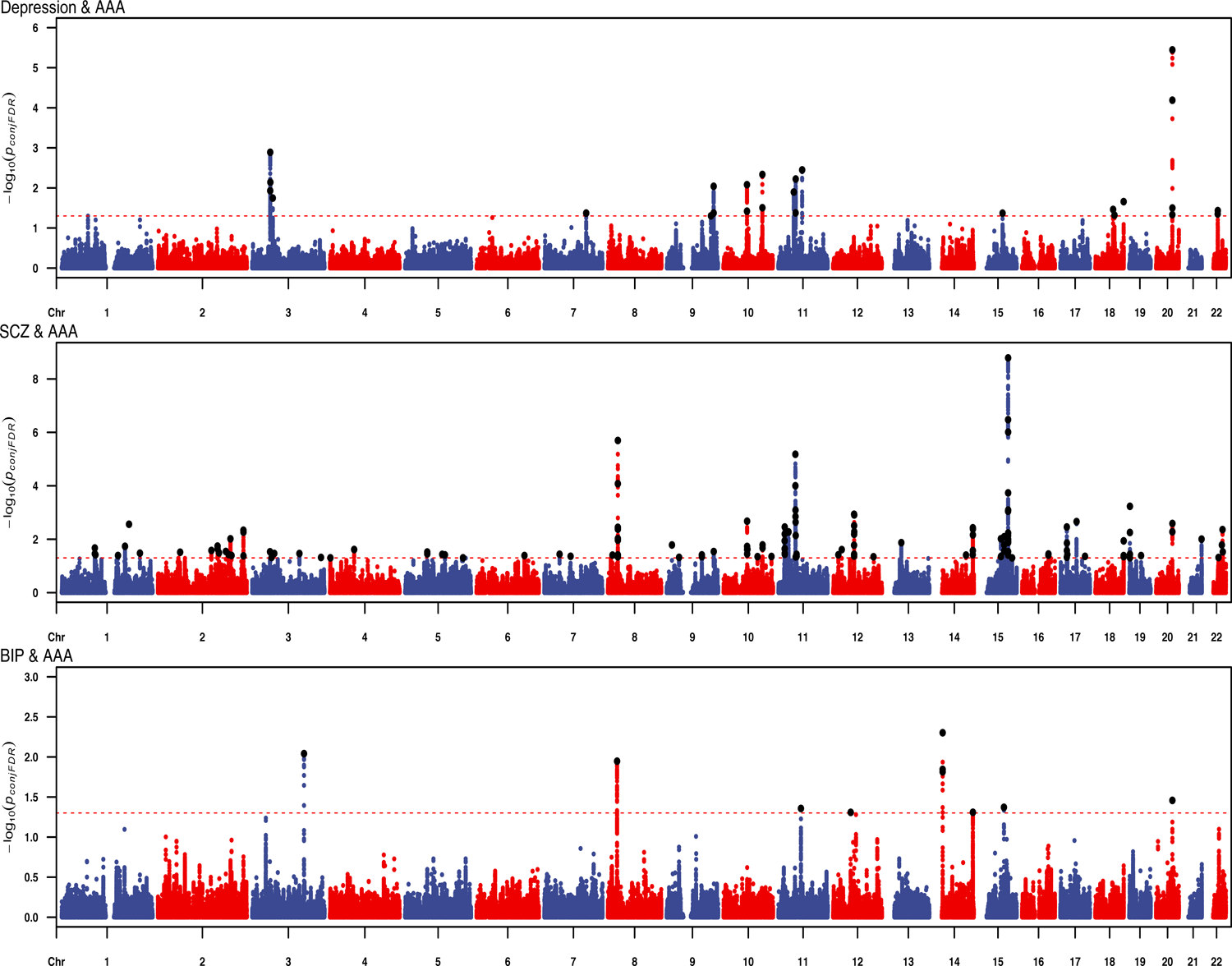
ConjFDR Manhattan plots. The red dashed line represents the threshold of *p*-value for conjFDR < 0.05, and the black points are the independent SNPs; SCZ, schizophrenia; BIP, bipolar disorder; AAA, abdominal aortic aneurysm.

### 3.2. Causal inference

#### 3.2.1. Univariate Mendelian Randomization

We initially examined the bidirectional causal relationship between mental disorders and AAA using the UMR, and the results are illustrated in **Fig. 4**. In the forward analyses, the MR-Egger regression intercept suggested the absence of horizontal pleiotropy among the selected SNPs (*p* values all > 0.05). The Cochran’s Q test revealed significant heterogeneity among the instruments concerning depression (Cochran’s Q = 163.448, *p* < 0.001), SCZ (Cochran’s Q = 333.578, *p* < 0.001), and BIP (Cochran’s Q =32.604, *p* = 0.002). Consequently, the primary causal inference utilized the random-effect IVW method. The IVW method revealed that people with depression (OR: 1.270, 95% CI: 1.071-1.504, *p* = 0.006) and SCZ (OR: 1.047, 95% CI: 1.010-1.084, *p* = 0.011) have a heightened susceptibility to AAA, while no causal effect of BIP on AAA was found (OR: 0.980, 95% CI: 0.896-1.121, *p* = 0.653). The findings mentioned above are supported by the weighted median, MR-CML, and MR-RAPS methods.

**Fig 4.**
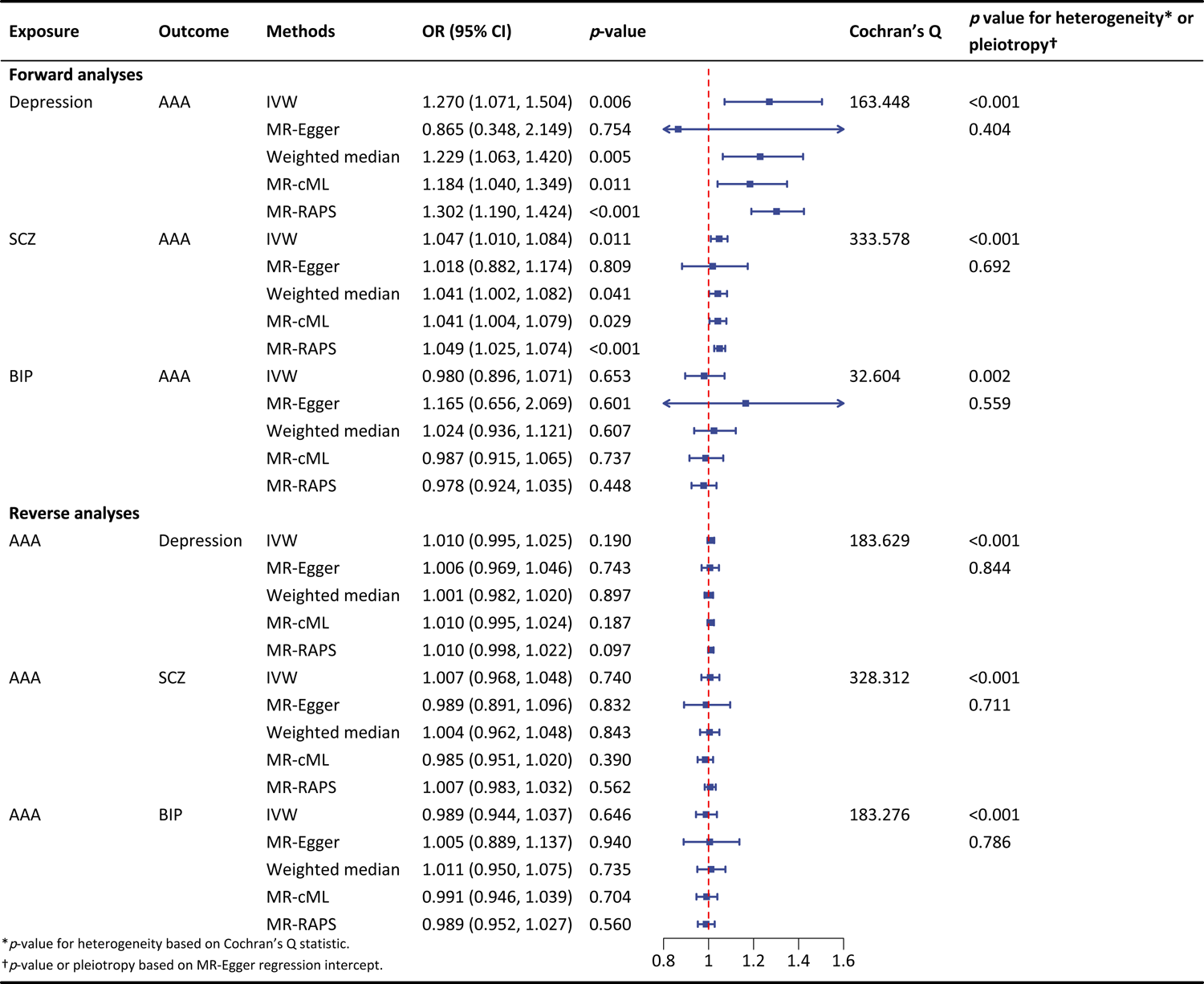
The univariate bidirectional causal inference between mental disorders and abdominal aortic aneurysm. MDD, major depression disorder; SCZ, schizophrenia; BIP, bipolar disorder; AAA, abdominal aortic aneurysm.

For the reverse analyses, the MR-Egger regression intercept also demonstrated there was no horizontal pleiotropy among the AAA-associated instrumental variables, and obvious heterogeneity was identified. The IVW method revealed that genetically predicted AAA showed no association with the risks of depression (OR: 1.010, 95% CI: 0.995-1.025, *p* = 0.190), SCZ (OR: 1.007, 95% CI: 0.968-1.048, *p* = 0.740), as well as BIP (OR: 0.989, 95% CI: 0.944-1.037, *p* = 0.646), and these null associations were consistently supported by estimates from the MR-Egger, weighted median, MR-cML, and MR-RAPS methods.

#### 3.2.2. Multivariate Mendelian Randomization

After performing the UMR, we proceeded with the MVMR to evaluate potential bias in the causal estimates due to confounding effects. The PhenoScanner revealed that instruments associated with mental disorders were predominantly linked to body mass index (BMI) **(Table S2)**. Additionally, SNPs associated with AAA were found to be correlated with BMI, coronary heart disease (CAD), high-density lipoprotein cholesterol (HDL-C), low-density lipoprotein cholesterol (LDL-C), and total cholesterol (TC) **(Table S3)**. Consequently, we conducted bidirectional MVMR to evaluate the multivariate-adjusted causal estimates between mental disorders and AAA. The forward analyses aimed to assess the effect of mental disorders on AAA with adjustments for BMI, while the reverse analyses aimed to evaluate the effect of AAA on mental disorders with adjustments for BMI, CAD, HDL-C, LDL-C, and TC.

The results of MVMR are presented in **Table S4**. The IVW method revealed that individuals with depression (OR: 1.315, 95% CI: 1.095-1.578, *p* = 0.003) and SCZ (OR: 1.042, 95% CI:1.0004-1.084, *p* = 0.048) were correlated with a higher risk of AAA even after controlling the effect of BMI, while people with BIP were not (OR: 1.014, 95% CI: 0.955-1.076, *p* = 0.652). Consistent with the results of UMR, the IVW methods also presented that genetically predicted AAA still showed no association with the risks of depression, SCZ, and BIP after adjustments for BMI, CAD, HDL-C, LDL-C, and TC. These bidirectional relationships were supported by the sensitivity analysis methods.

#### 3.2.3. Mediation Mendelian Randomization

Since the UMR and MVMR only supported the causal effects of depression and SCZ on AAA, we would only investigate whether these two causalities were mediated by the known risk factors of AAA. As described in the methods section, we selected smoking, hypertension, hyperlipidemia, and CA as the four candidate mediating factors.

The results of mediation MR analyses are presented in **Table S5**. The first step MR analyses presented that genetically predicted depression was associated with increased odds ratio of smoking (OR: 1.236, 95% CI: 1.140-1.340, *p* <0.001), hypertension (OR: 1.206, 95% CI: 1.069-1.361, *p* = 0.002), hyperlipidemia (OR: 1.200, 95% CI: 1.037-1.388, *p* = 0.015), and CA (OR: 1.210, 95% CI: 1.078-1.359, *p* = 0.001). In contrast, genetically predicted SCZ was only associated with an increased likelihood of smoking (OR: 1.047, 95% CI: 1.023-1.071, *p* <0.001). The second step MR analyses demonstrated that genetically predicted smoking (OR: 1.610, 95% CI: 1.449-1.788, *p* <0.001), hypertension (OR: 1.218, 95% CI: 1.142-1.300, *p* <0.001), hyperlipidemia (OR: 1.547, 1.288-1.857, *p* <0.001), and CA (OR: 1.385, 95% CI: 1.204-1.594, *p* <0.001) were all associated with elevated risks of AAA. Based on the results of mediation MR findings, the causal effect of depression on AAA was mediated by smoking, hypertension, hyperlipidemia, and CA, while the causal effect of SCZ on AAA was mediated by smoking only (**Fig 5**).

**Fig 5.**
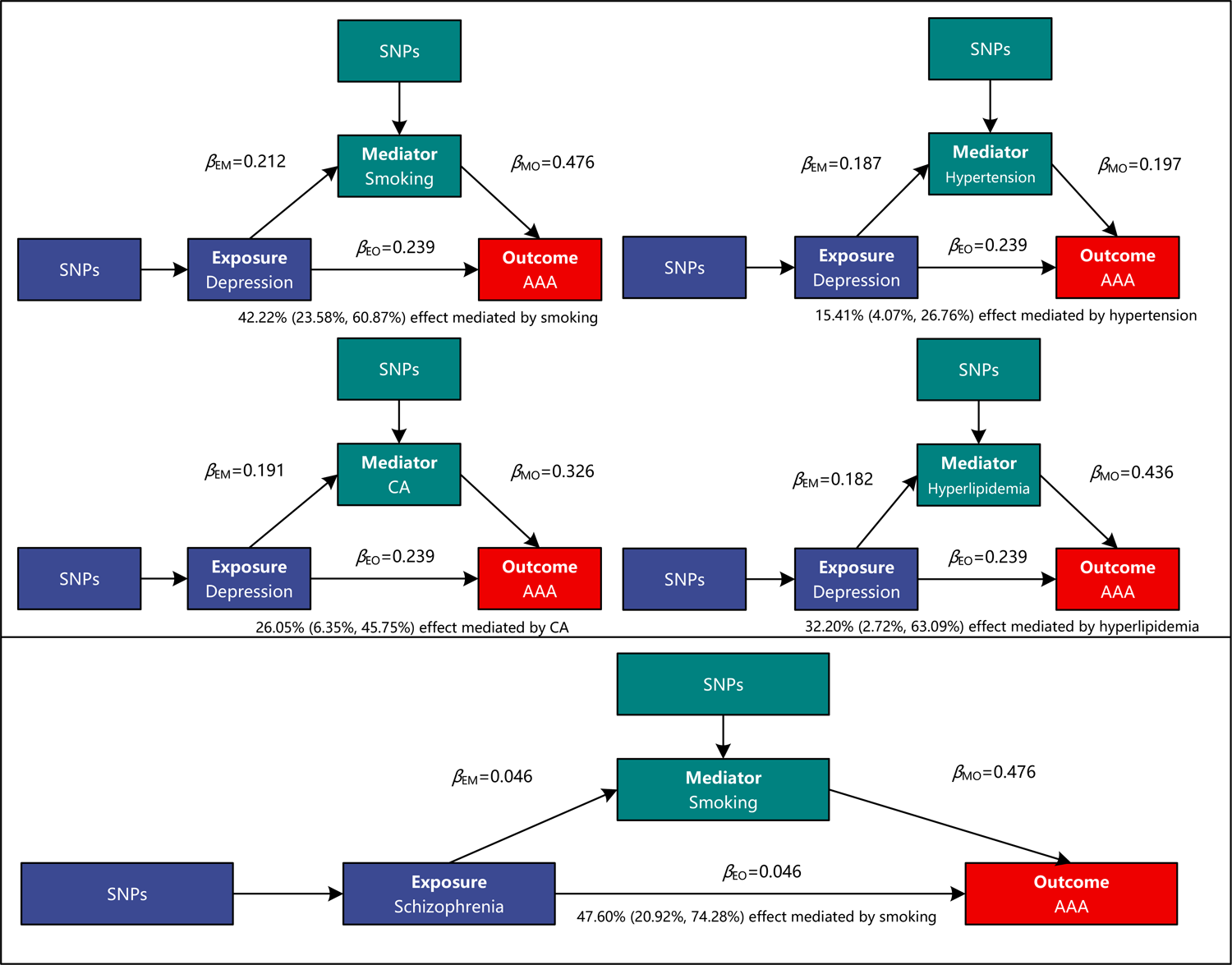
Mediation effects of risk factors in the causal associations of abdominal aortic aneurysm with depression and schizophrenia. SNPs, single nucleotide polymorphisms; AAA, abdominal aortic aneurysm; CA, coronary atherosclerosis.

## 4. Discussion

We conducted an investigation into the relationship between mental disorders and AAA from the genetic perspective. Our findings provided solid evidence of the genetic correlation between AAA and depression as well as SCZ. Besides, the causal effects of depression and SCZ on AAA were identified, with known risk factors for AAA playing a mediating role.

Few studies have investigated the associations between mental disorders and AAA, with the evidence supporting the correlations between depression and AAA being limited. Furthermore, a study involving 574 participants from four centers in Australia suggested an insignificant association between depression and AAA growth^25^, contradicting conclusions drawn in previous studies from another perspective. Given the scarcity of research and the inconsistency of results, we conducted comprehensive genetic association, and univariate, multivariate as well as mediating MR to evaluate the correlation and causal links between mental disorders and AAA.

The global genetic correlation analyses indicated positive associations of AAA with depression and SCZ, and meanwhile, numerous locally correlated genomic regions and shared loci were identified. The genetic association analyses presented that AAA had plenty of genetic basis with mental disorders. The results of univariate and multivariate MR analyses demonstrated robust and statistically significant associations of depression, SCZ, and AAA, and we further extended our investigation through mediation analysis to delineate potential pathways underlying these associations. By incorporating well-established risk factors of AAA as potential mediators, significant mediation effects of smoking, hyperlipidemia, CA, and hypertension were identified in the causal pathways between depression and AAA, and the proportions of mediation effects were calculated as 42.22%, 32.20%, 26.05%, and 15.41%, respectively. In addition, smoking exhibited a substantial mediation effect proportion of 47.60% in the association between SCZ and AAA. These findings underscore the pivotal role of smoking in the nexus between mental disorders and AAA, offering crucial insights for further etiological explorations.

While the pathological mechanism of AAA remains elusive, existing literature has elucidated a series of contributing processes, notably the infiltration and subsequent destruction of the aortic wall mediated by immune and inflammatory mechanisms^26^. Both innate and adaptive immune cells, including macrophages and T cells, have undergone extensive scrutiny in the pathogenesis of AAA, with several inflammasomes suggested to be involved in both the inhibition and amelioration of AAA^27,28^. NF-κB, the primary regulator of inflammation and immune response, is activated by multiple factors including angiotensin II, triggering macrophage infiltration, inflammation, and subsequent aortic deterioration^29^. Numerous studies have explored the impact of pharmacological inhibition of nucleotide-binding oligomerization domain-like receptor family protein 3 (NLRP3) inflammasome on experimental AAAs, revealing preventive effects characterized by reduced AAA diameter and incidence^30–32^, etc. Furthermore, the remarkable parallels between the immune system and the brain, both intricately integrated and complex systems associated with memory, imply intimate connections between immune responses and mental disorders. For instance, some researchers consider depression as an inflammatory disease^33,34^, with inflammation likely playing a critical role in heightening individual susceptibility to depression^35^. Similarly, extensive reviews have highlighted the central role of inflammation and immunity in SCZ, findings supported by both human and animal studies^36,37^. Several two-sample MR studies indicate the causal associations of inflammatory biomarkers such as C-reactive protein and soluble interleukin-6 receptor with SCZ, with anti-inflammation strategies emerging as therapeutic considerations^38,39^. Therefore, inflammation and immune responses may function as a conduit linking mental disorders and AAA.

Similar to other cardiovascular diseases (CVD), the relationship between mental disorders and AAA may also be explained by the shared etiological factors, encompassing genetic, biological, and behavioral mechanisms. Early in 2002, Bondy et al. found that the same allelic combination of two genes could increase the vulnerability for depressive disorder while also being associated with a higher risk of myocardial infarction^40^, suggesting the genetic associations between depression and CVD. Molecular genetic studies have further corroborated modest genetic correlations between severe mental illness and CVD^41^.

Furthermore, biological constituents such as neutrophil gelatinase associated lipocalin^42^ and physiological processes like platelet receptors^43^ and function have been implicated as mediating factors linking CVD and depression. Moreover, the overlap of behavioral determinants including smoking, physical inactivity, obesity, and poor diet was observed among participants affected by mental disorders and CVD. Similarly, GWAS studies have unveiled extensive genetic convergence between SCZ and CVD risk factors, with notable associations observed with smoking initiation and body mass index^44^.

Our study had several strengths worth pointing out. Firstly, we utilized the genetic data from the large-scale GWASs, which could significantly enhance the statistical power and avoid the confounding bias and reverse causality of observational studies. Additionally, we conducted two parts of analyses, including genetic correlation and causality inference, and the results provided consistent evidence on the causal effects of depression and SCZ on AAA. Furthermore, we identified the mediation effect, which highlighted the interactive impacts of mental disorders and other risk factors on the risk of AAA.

Despite these advantages, our findings should be interpreted in the context of limitations. Firstly, the most recent GWAS of BIP have identified various significant SNPs^45^, but the samples overlapped with the AAA. To avoid the causal inference bias due to sample overlap, we selected the genetic variants associated with BIP from the GWAS with relatively lower sample sizes^13^. Further studies were warranted to check the relationship between BIP and AAA. In addition, only the summary-level data could be obtained thus making us unable to perform the stratified analyses. At the same time, current knowledge indicates that the prevalence of AAA in males was higher than in females. Further research was needed to confirm our findings in specific sub-populations. Lastly, the samples were all of European ancestry, making our findings may not apply to other populations.

## 5. Conclusion

Genetically predicted depression and schizophrenia were associated with elevated risk of abdominal aortic aneurysm, and the known risk factors of abdominal aortic aneurysm played mediation roles.

## 6. Declarations

## 6.1. Acknowledgement

None.

## 6.2. Ethical approval

The datasets used in this study were all publicly available from previous studies, and the ethical approvals were obtained by the original researchers. Therefore, the ethical approval for this study could be waived.

## 6.3. Consent for publication

Not applicable

## 6.4. Data availability

The datasets utilized in this study are publicly available; the details can be found in the supplementary method.

## 6.5. Code availability

ldscr (https://github.com/bulik/ldsc), GNOVA (https://github.com/xtonyjiang/GNOVA), SU PERGNOVA (https://github.com/qlu-lab/SUPERGNOVA), pleioFDR (https://github.com/pr ecimed/pleiofdr), TwoSampleMR (https://mrcieu.github.io/TwoSampleMR/), MendelianRa ndomization (https://github.com/cran/MendelianRandomization), and mr.raps (https://githu b.com/qingyuanzhao/mr.raps).

## 6.8. Author contributions

**Ming-Gang Deng:** Conceptualization, Methodology, Data Curation, Formal Analysis, and Writing - Original Draft; **Chen Chai:** Writing - Original Draft; Kai Wang: Writing - Original Draft; **Zhi-Hui Zhao:** Writing – review & editing; **Jia-Qi Nie**: Writing – review & editing; **Fang Liu**: Writing – review & editing; **Yuehui Liang:** Writing – review & editing; **Jiewei Liu:** Conceptualization, and Writing - Original Draft.

## Reference

1. Summers KL, Kerut EK, Sheahan CM, Sheahan MG, 3rd. Evaluating the prevalence of abdominal aortic aneurysms in the United States through a national screening database. Journal of vascular surgery. 2021;73(1):61–68.

2. Davis FM, Rateri DL, Daugherty A. Abdominal aortic aneurysm: novel mechanisms and therapies. Current opinion in cardiology. 2015;30(6):566–573.

3. Zhang F, Li K, Zhang W, et al. Ganglioside GM3 Protects Against Abdominal Aortic Aneurysm by Suppressing Ferroptosis in Vascular Smooth Muscle Cells. Circulation. 2023.

4. Deng MG, Liu F, Liang Y, Wang K, Nie JQ, Liu J. Association between frailty and depression: A bidirectional Mendelian randomization study. Science advances. 2023;9(38):eadi3902.

5. Deng M-G, Wang K, Liu F, Zhou X, Liu J. The Relationship Between Frailty and Schizophrenia: A Genetic Association and Mendelian Randomization Study. 2023.

6. Li GH, Cheung CL, Chung AK, et al. Evaluation of bi-directional causal association between depression and cardiovascular diseases: a Mendelian randomization study. Psychological medicine. 2022;52(9):1765–1776.

7. Veeneman RR, Vermeulen JM, Abdellaoui A, et al. Exploring the Relationship Between Schizophrenia and Cardiovascular Disease: A Genetic Correlation and Multivariable Mendelian Randomization Study. Schizophrenia bulletin. 2022;48(2):463–473.

8. So HC, Chau KL, Ao FK, Mo CH, Sham PC. Exploring shared genetic bases and causal relationships of schizophrenia and bipolar disorder with 28 cardiovascular and metabolic traits. Psychological medicine. 2019;49(8):1286–1298.

9. Kim MH, Yoo JH, Cho HJ, et al. Increased depression risk in patients with abdominal aortic aneurysm: a nationwide cohort study. Annals of surgical treatment and research. 2021;101(5):291–298.

10. Nyrønning L, Stenman M, Hultgren R, Albrektsen G, Videm V, Mattsson E. Symptoms of Depression and Risk of Abdominal Aortic Aneurysm: A HUNT Study. Journal of the American Heart Association. 2019;8(21):e012535.

11. Howard DM, Adams MJ, Clarke TK, et al. Genome-wide meta-analysis of depression identifies 102 independent variants and highlights the importance of the prefrontal brain regions. Nature neuroscience. 2019;22(3):343–352.

12. Trubetskoy V, Pardiñas AF, Qi T, et al. Mapping genomic loci implicates genes and synaptic biology in schizophrenia. Nature. 2022;604(7906):502–508.

13. Stahl EA, Breen G, Forstner AJ, et al. Genome-wide association study identifies 30 loci associated with bipolar disorder. Nature genetics. 2019;51(5):793–803.

14. Roychowdhury T, Klarin D, Levin MG, et al. Genome-wide association meta-analysis identifies risk loci for abdominal aortic aneurysm and highlights PCSK9 as a therapeutic target. Nature genetics. 2023;55(11):1831–1842.

15. Emdin CA, Khera AV, Kathiresan S. Mendelian Randomization. Jama. 2017;318(19):1925–1926.

16. Deng MG, Cui HT, Nie JQ, Liang Y, Chai C. Genetic association between circulating selenium level and the risk of schizophrenia in the European population: A two-sample Mendelian randomization study. Frontiers in nutrition. 2022;9:969887.

17. Deng MG, Liu F, Wang K, Liang Y, Nie JQ, Chai C. Genetic association between coffee/caffeine consumption and the risk of obstructive sleep apnea in the European population: a two-sample Mendelian randomization study. European journal of nutrition. 2023;62(8):3423–3431.

18. Yu X, Deng MG, Tang ZY, Zhang ZJ. Urticaria and increased risk of rheumatoid arthritis: a two-sample Mendelian randomisation study in European population. Modern rheumatology. 2022;32(4):736–740.

19. Bulik-Sullivan B, Finucane HK, Anttila V, et al. An atlas of genetic correlations across human diseases and traits. Nature genetics. 2015;47(11):1236–1241.

20. Lu Q, Li B, Ou D, et al. A Powerful Approach to Estimating Annotation-Stratified Genetic Covariance via GWAS Summary Statistics. American journal of human genetics. 2017;101(6):939–964.

21. Zhang Y, Lu Q, Ye Y, et al. SUPERGNOVA: local genetic correlation analysis reveals heterogeneous etiologic sharing of complex traits. Genome biology. 2021;22(1):262.

22. Andreassen OA, Thompson WK, Schork AJ, et al. Improved detection of common variants associated with schizophrenia and bipolar disorder using pleiotropy-informed conditional false discovery rate. PLoS genetics. 2013;9(4):e1003455.

23. Sanderson E. Multivariable Mendelian Randomization and Mediation. Cold Spring Harbor perspectives in medicine. 2021;11(2).

24. Kamat MA, Blackshaw JA, Young R, et al. PhenoScanner V2: an expanded tool for searching human genotype-phenotype associations. Bioinformatics (Oxford, England). 2019;35(22):4851–4853.

25. Thanigaimani S, Phie J, Quigley F, et al. Association of Diagnosis of Depression and Small Abdominal Aortic Aneurysm Growth. Annals of vascular surgery. 2022;79:256–263.

26. Márquez-Sánchez AC, Koltsova EK. Immune and inflammatory mechanisms of abdominal aortic aneurysm. Frontiers in immunology. 2022;13:989933.

27. Yuan Z, Lu Y, Wei J, Wu J, Yang J, Cai Z. Abdominal Aortic Aneurysm: Roles of Inflammatory Cells. Frontiers in immunology. 2020;11:609161.

28. Raffort J, Lareyre F, Clément M, Hassen-Khodja R, Chinetti G, Mallat Z. Monocytes and macrophages in abdominal aortic aneurysm. Nature reviews Cardiology. 2017;14(8):457–471.

29. Li Z, Kong W. Cellular signaling in Abdominal Aortic Aneurysm. Cellular signalling. 2020;70:109575.

30. Tsuruda T, Kato J, Hatakeyama K, et al. Adventitial mast cells contribute to pathogenesis in the progression of abdominal aortic aneurysm. Circulation research. 2008;102(11):1368–1377.

31. Yamanouchi D, Morgan S, Kato K, Lengfeld J, Zhang F, Liu B. Effects of caspase inhibitor on angiotensin II-induced abdominal aortic aneurysm in apolipoprotein E-deficient mice. Arteriosclerosis, thrombosis, and vascular biology. 2010;30(4):702–707.

32. Kaneko H, Anzai T, Morisawa M, et al. Resveratrol prevents the development of abdominal aortic aneurysm through attenuation of inflammation, oxidative stress, and neovascularization. Atherosclerosis. 2011;217(2):350–357.

33. Berk M, Williams LJ, Jacka FN, et al. So depression is an inflammatory disease, but where does the inflammation come from? BMC medicine. 2013;11:200.

34. Halaris A. Inflammation and depression but where does the inflammation come from? Current opinion in psychiatry. 2019;32(5):422–428.

35. Beurel E, Toups M, Nemeroff CB. The Bidirectional Relationship of Depression and Inflammation: Double Trouble. Neuron. 2020;107(2):234–256.

36. Müller N. Inflammation in Schizophrenia: Pathogenetic Aspects and Therapeutic Considerations. Schizophrenia bulletin. 2018;44(5):973–982.

37. Khandaker GM, Cousins L, Deakin J, Lennox BR, Yolken R, Jones PB. Inflammation and immunity in schizophrenia: implications for pathophysiology and treatment. The lancet Psychiatry. 2015;2(3):258–270.

38. Hartwig FP, Borges MC, Horta BL, Bowden J, Davey Smith G. Inflammatory Biomarkers and Risk of Schizophrenia: A 2-Sample Mendelian Randomization Study. JAMA psychiatry. 2017;74(12):1226–1233.

39. Williams JA, Burgess S, Suckling J, et al. Inflammation and Brain Structure in Schizophrenia and Other Neuropsychiatric Disorders: A Mendelian Randomization Study. JAMA psychiatry. 2022;79(5):498–507.

40. Bondy B, Baghai TC, Zill P, et al. Combined action of the ACE D- and the G-protein beta3 T-allele in major depression: a possible link to cardiovascular disease? Molecular psychiatry. 2002;7(10):1120–1126.

41. Amare AT, Schubert KO, Klingler-Hoffmann M, Cohen-Woods S, Baune BT. The genetic overlap between mood disorders and cardiometabolic diseases: a systematic review of genome wide and candidate gene studies. Translational psychiatry. 2017;7(1):e1007.

42. Gouweleeuw L, Naudé PJ, Rots M, DeJongste MJ, Eisel UL, Schoemaker RG. The role of neutrophil gelatinase associated lipocalin (NGAL) as biological constituent linking depression and cardiovascular disease. Brain, behavior, and immunity. 2015;46:23–32.

43. Amadio P, Zarà M, Sandrini L, Ieraci A, Barbieri SS. Depression and Cardiovascular Disease: The Viewpoint of Platelets. International journal of molecular sciences. 2020;21(20).

44. Rødevand L, Rahman Z, Hindley GFL, et al. Characterizing the Shared Genetic Underpinnings of Schizophrenia and Cardiovascular Disease Risk Factors. The American journal of psychiatry. 2023;180(11):815–826.

45. Mullins N, Forstner AJ, O’Connell KS, et al. Genome-wide association study of more than 40,000 bipolar disorder cases provides new insights into the underlying biology. Nature genetics. 2021;53(6):817–829.

