## Supplementary Method for "Causal relationship between mental disorders and abdominal aortic aneurysm: insights from the genetic perspective"

The genetic variants associated with AAA were extracted from a meta-analysis of 17 individual GWASs from 14 cohorts in the AAAgen Consortium, which included 1,125,328 participants and were primarily from 4 cohorts: Copenhagen Hospital Biobank-Cardiovascular Disease Cohort and Danish Blood Donor Study (CHB-CVDC+DBDS), deCODE, DiscovEHR, and Million Veteran Program (MVP)^1^. The meta-analysis identified a total of 141 independent associations, 97 of which were previously unreported loci.

The genetic data associated with depression was obtained from the recent genome-wide meta-analysis of 246,363 cases and 561,190 controls from three GWASs of depression^2^, which included 307,354 samples from 23andMe, 361,315 individuals from the UK Biobank, and 138,884 participants from the Psychiatric Genomics Consortium (PGC)-2018^3^. Due to the data accessibility, we excluded people from 23andMe, resulting in a sub-sample of 170,756 cases and 329,443 controls.

The genetic data related to schizophrenia (SCZ) were obtained from the GWAS of up to 76,755 individuals with SCZ and 243,649 control individuals, involving samples from individuals of European, East Asian, African American, and Latino ancestry^4^. We extracted the European ancestry-specific data, with a sub-sample of 52,017 individuals with schizophrenia and 75,889 controls.

The most recent GWAS of bipolar disorder (BIP) identified 64 significant genomic loci in 41,917 cases and 371,549 controls of European ancestry^5^. However, the majority of the samples (47.06%) came from the deCODE, which overlaps with the samples of AAA. To mitigate the bias of sample overlap, we obtained the BIP-associated genetic data from a previous GWAS, which included 20,352 cases and 31,358 controls of European descent^6^.

The details for the genetic data used for multivariate Mendelian Randomization (MR) and mediation MR analyses are presented in the table below.

In general, we selected the data sources for these phenotypes considering European ancestry specificity, a larger sample size, and a lower sample overlap. A larger sample size enhances the statistical power, and a lower sample overlap avoids bias in causal inference, especially for the two-sample MR analyses^7^.

| Phenotype | PMID | Sample size (overall or case/control) | Ancestry | Consortium |
| --- | --- | --- | --- | --- |
| Data sources for AAA and mental disorders | | | | |
| AAA | 37845353 | 392,21/1,086,107 | European | [AAAgen](https://csg.sph.umich.edu/willer/public/AAAgen2023/) |
| Depression | 30718901 | 170,756/329,443 | European | [Psychiatric Genomics Consortium](https://pgc.unc.edu/for-researchers/download-results/) |
| Schizophrenia | 35396580 | 52,017/75,889 | European | [Psychiatric Genomics Consortium](https://pgc.unc.edu/for-researchers/download-results/) |
| Bipolar disorder | 31043756 | 20,352/31,358 | European | [Psychiatric Genomics Consortium](https://pgc.unc.edu/for-researchers/download-results/) |
| Data sources for covariates used in the multivariate Mendelian Randomization | | | | |
| BMI | 25673413 | 322,154 | European | [The Genetic Investigation of ANthropometric Traits](https://portals.broadinstitute.org/collaboration/giant/index.php/GIANT_consortium) |
| CAD | 26343387 | 60,801/123,504 | European | [CARDIoGRAMplusC4D](https://gwas.mrcieu.ac.uk/datasets/ieu-a-7/) |
| TC | 34887591 | 930,672 | European | [The Global Lipids Genetics Consortium](https://csg.sph.umich.edu/willer/public/glgc-lipids2021/results/ancestry_specific/) |
| HDL-C | 34887591 | 888,227 | European | [The Global Lipids Genetics Consortium](https://csg.sph.umich.edu/willer/public/glgc-lipids2021/results/ancestry_specific/) |
| LDL-C | 34887591 | 842,660 | European | [The Global Lipids Genetics Consortium](https://csg.sph.umich.edu/willer/public/glgc-lipids2021/results/ancestry_specific/) |
| Data sources for candidate mediators used in the mediation Mendelian Randomization | | | | |
| Smoking | 30643251 | 311,629/321,173 | European | [GSCAN Consortium](https://gwas.mrcieu.ac.uk/datasets/ieu-b-4877/) |
| Hypertension | NA | 111,581/265,626 | European | [FinnGen](https://r9.finngen.fi/) |
| Hyperlipidemia | NA | 12,304/324,150 | European | [FinnGen](https://r9.finngen.fi/) |
| CA | NA | 47,550/313,400 | European | [FinnGen](https://r9.finngen.fi/) |

**References**

1.	Roychowdhury T, Klarin D, Levin MG, et al. Genome-wide association meta-analysis identifies risk loci for abdominal aortic aneurysm and highlights PCSK9 as a therapeutic target. *Nature genetics.* 2023;55(11):1831-1842.

2.	Howard DM, Adams MJ, Clarke TK, et al. Genome-wide meta-analysis of depression identifies 102 independent variants and highlights the importance of the prefrontal brain regions. *Nature neuroscience.* 2019;22(3):343-352.

3.	Wray NR, Ripke S, Mattheisen M, et al. Genome-wide association analyses identify 44 risk variants and refine the genetic architecture of major depression. *Nature genetics.* 2018;50(5):668-681.

4.	Trubetskoy V, Pardiñas AF, Qi T, et al. Mapping genomic loci implicates genes and synaptic biology in schizophrenia. *Nature.* 2022;604(7906):502-508.

5.	Mullins N, Forstner AJ, O'Connell KS, et al. Genome-wide association study of more than 40,000 bipolar disorder cases provides new insights into the underlying biology. *Nature genetics.* 2021;53(6):817-829.

6.	Stahl EA, Breen G, Forstner AJ, et al. Genome-wide association study identifies 30 loci associated with bipolar disorder. *Nature genetics.* 2019;51(5):793-803.

7.	Burgess S, Davies NM, Thompson SG. Bias due to participant overlap in two-sample Mendelian randomization. *Genetic epidemiology.* 2016;40(7):597-608.
